# Discovering age- and sex-specific genetic risk factors in sensorineural hearing loss: genome-wide evidence from large-scale biobank studies

**DOI:** 10.1101/2025.05.12.25327483

**Authors:** Argyro Bizaki-Vallaskangas, Eeva Sliz, Tuuli Lankinen, Elmo Saarentaus, Ville Salo, Kristi Krebs, Tytti Willberg, Ilkka Kivekäs, Joel Rämö, Sanna Toppila-Salmi, Aarno Dietz, Vesa Hytönen, Aarno Palotie, Lili Milani, Antti Mäkitie, Johannes Kettunen

## Abstract

**Purpose:** Investigate the genetic components of sensorineural hearing loss (SNHL) by performing genome-wide meta-analyses using the data from FinnGen and Estonian Biobank.

**Methods:** We studied genome-wide associations of SNHL in FinnGen and the Estonian Biobank in the general population and in sex- and age-of-onset stratified subgroups. The study-specific GWASs were combined through inverse variance-weighted genome-wide meta-analyses, encompassing a total of 531,059 individuals (Ncases=35,960). Age-stratified meta-analyses included 28,198 individuals diagnosed at the age of 55 years or after and 7,762 individuals diagnosed before the age of 55 years, with 495,099 controls. Sex-stratified meta-analyses included 313,501 females (Ncases=17,761) and 217,558 males (Ncases=18,199).

**Results:** In the meta-analysis focusing on the general population, 22 SNHL-associated loci (±1 Mb window) were observed, seven of which were previously unreported. In the sex-stratified analysis, two previously unreported SNHL loci were observed in the female subgroup and one locus in the male subgroup. Additionally, in the age-stratified analysis, three previously unreported SNHL loci were observed in the subgroup of those that were diagnosed at the age of 55 years or after. In those diagnosed before the age of 55 years, one previously unreported locus was observed. Overall, 32 loci were associated with SNHL at p<5×10^−8^ in at least one of the study groups. Of these, 14 loci have not been previously reported in association with SNHL. We also estimated if there were significant differences in the effect sizes of the lead variants at each locus between the analytical subgroups and observed differences for 14 variants.

**Conclusions:** Previously unreported SNHL risk loci and differences in effect sizes found in this study provide additional insight into the genetic underpinnings of SNHL. Our results validate the role of mechano-transduction and genetic components affecting the structure of the inner ear in the background of SNHL. Our study contributes to our understanding of the genetic causes of SNHL and may open the door for further research into innovative therapies and preventative measures.

## Introduction

Sensorineural hearing loss is the most common sensory deficit affecting individuals across all ages. It may have genetic or environmental origins and can present as either syndromic, occurring alongside other clinical features, or non-syndromic, where hearing loss is the sole manifestation. Sensorineural hearing loss (SNHL) results from damage to the hair cells of the inner ear or the vestibulocochlear nerve. While hair cell damage is a major contributor to SNHL, there are also other mechanisms such as synaptopathy or stria vascularis dysfunctions, that may lead to SNHL.Presbyacusis, or age-related hearing loss (ARHL), is the most prevalent form of hearing loss, which affects approximately 35% of those aged 65–74 years and 50% of those aged 75 and older ^1,2,56^.

It is usually characterized by progressive bilateral high-frequency hearing loss, which impairs speech recognition, especially in noisy environments^3–5^. The quality of life can be severely impacted by hearing loss, necessitating significant changes to daily routines. Also, SHNL is a major disability that also causes significant economic costs^28,29^. It is also linked to severe comorbidities, such as dementia, depression, and social isolation, all of which can further worsen quality of life^6,7,57^.

The heritability for hearing loss is estimated to be between 0.36-0.75 based on twin and familial studies^8–11^, indicating that genetic elements are an important contributor to the risk of SNHL. Previous genome-wide association studies (GWASs) have identified numerous loci associated with SNHL and defined the complex nature of SNHL^12–26^. Due to varying definitions of the phenotypes, studies in isolated populations, and overall modest sample sizes, some of these earlier GWASs have lacked consistency Because of these factors and the high degree of genetic heterogeneity in SNHL, the findings have not been consistently replicated in following studies^13^. Clinical studies have shown that a patient’s sex might affect the presentation time and severity of SNHL27, but the underlying genetic basis is still poorly understood. Despite the high prevalence of SNHL in the ageing population, only a few age-stratified GWASs have been conducted.

Due to the shortcomings, the present study was designed to identify age of onset or sex-specific loci with stratified GWAS analyses. Here, we present the results of genome-wide meta-analyses of SNHL using the data from FinnGen and the Estonian Biobank. We were able to find novel loci in the combined meta-analysis and to demonstrate that stratifying SNHL by sex and age provides additional loci, and genetic variants exhibit differential effect estimates depending on the stratification of research participants.

## Materials & Methods

### Study populations

**FinnGen** is a collaborative project launched in 2017, aiming at advancing human health through genetic research. This initiative harnesses genome data collected from a nationwide network of Finnish biobanks, which are linked with digital health records from various national registries, including hospital discharge, death, cancer, and medication reimbursement databases. The project covers approximately 10% of the Finnish population. FinnGen Data Freeze 9 data utilized in the study included 27,232 SHNL cases and 326,172 controls (Table S1).

The Estonian Biobank (EstBB) is a longitudinal biobank in Northern Europe that holds genotype and phenotype data for 20% of the Estonian adult population ^30,58^. At recruitment, participants signed a consent allowing follow-up linkage of their electronic health records (EHR). Health records at EstBB are extracted from the data provided by regional hospitals, national registries, and the Estonian Health Insurance Fund, a national administrative database that pools detailed and individual-level billing data for all health care services as well as digital prescription information. The activities of the EstBB are regulated by the Human Genes Research Act, which was adopted in 2000 specifically for the operations of the EstBB. Individual-level data analysis in the EstBB was carried out under ethical approval number 1.1-12/624 from the Estonian Committee on Bioethics and Human Research (Estonian Ministry of Social Affairs). In this study, 177 655 participants from EstBB were included, 8728 cases and 168 927 controls (Table S1).

### Phenotype descriptions

We identified SNHL cases that were required to have an entry of *The International Classification of Diseases, Tenth Revision* (ICD-10): H90.3, H90.4, H90.5, H91.1. Participants without a record of these ICD codes were classified as controls. In addition, the individuals with the records of the following codes were excluded from the analyses: H65.4, H66.1, H66.2, H66.3, H67.0*A18.6, H67.0*A38, H67.1*, H67.1*B05.3, H68.0, H68.1, H70.0, H70.1, H70.2, H70.8, H70.9, H71, H73.1, H73.8, H73.9, H74.0, H74.1, H74.2, H74.3, H74.4, H74.8, H74.9, H75.0*, H75.0*A18.0, H75.8*, H80.0, H80.1, H80.20, H80.28, H80.8, H80.9, H81.0, H83.0, H83.1, H83.2, H83.3, H83.8, H83.9, H90.0, H90.1, H90.2, H90.6, H90.7, H90.8, H91.0#, H91.2, H91.3, H91.8, H91.9, H93.3, H93.8, H93.9, H94.0*, H94.0*A52.1, H94.8*, H95.0, H95.1, H95.8, H95.9, Q16.0, Q16.1, Q16.2, Q16.3, Q16.4, Q16.5, Q16.9, Q17.2, Q17.3, Q17.4, Q17.8, Q17.9, D33.30. Overall, the meta-analysis focusing on the general population consisted of 35,960 SNHL cases and 495,099 controls (Table S1).

In the sex-stratified analyses, participants were stratified to males (18,199 cases in the meta-analysis) and females (17,761 cases in the meta-analysis) and were compared to same-sex controls (the respective numbers of controls were 199,359 and 295,740). In the age-stratified analyses, the cases were classified into two groups: those who were diagnosed before the age of 55 (7,762 cases) and those who were diagnosed at the age of 55 or after (28,198 cases). The control group was not stratified by age in the age-stratified analyses (N_controls_=495,099 in the age-of-onset-stratified analysis). The number of participants in each studied cohort is available in Table S1.

### Genotyping, imputation & quality control

In FinnGen, genotyping of the samples was performed using Illumina and Affymetrix arrays (Illumina Inc., San Diego, and Thermo Fisher Scientific, Santa Clara, CA, USA). Sample quality control (QC) was performed to exclude individuals with high genotype missingness (>5%), ambiguous gender, excess heterozygosity (±4SD) and non-Finnish ancestry. Regarding variant QC, all variants with low Hardy-Weinberg equilibrium (HWE) p-value (<1e-6), high missingness (>2%) and minor allele count (MAC)<3 were excluded. Chip genotyped samples were pre-phased with Eagle 2.3.5, with the number of conditioning haplotypes set to 20,000. Genotype imputation was conducted by using the Finnish population-specific SISu v3 reference panel with Beagle 4.1 (version 08Jun17.d8b) as described in the following protocol: dx.doi.org/10.17504/protocols.io.nmndc5e. In post-imputation QC, variants with imputation INFO<0.6 were excluded.

Genotyping of the samples from the Estonian Biobank was conducted in the Core Genotyping Lab of the Institute of Genomics, University of Tartu, using the following Illumina arrays: GSAMD-24v1, GSAMD-24v2, ESTchip-1_GSAv2-MD, and ESTchip-2_GSAv3-MD. Quality control was conducted according to best practices (exclusion of individuals with call rate <95%, with a mismatch between genotype and phenotype sex and who deviated +/- 3SD from the samples heterozygosity rate mean; exclusion of SNVs with call rate <95%, HWE p<1e-4, MAF <1%). Samples were pre-phased with Eagle v2.3 ^59^ with the number of conditioning haplotypes set to 20,000, and genotype imputation was carried out with Beagle v.28Sep18.793 using an Estonian-specific reference panel^31,60^. Prior to imputation, variants with minor allele frequency (MAF)<1% and indels were removed. Further, EstBB samples were combined with the 1000 genomes phase 3 dataset for ancestry analysis. Genetic principal components were calculated using a subset of quality-controlled and pruned genotyped SNPs. This was further used to identify and remove samples that deviated from the main cluster.

### GWASs and meta-analyses

GWASs were conducted with REGENIE (v2.2.4) using an additive model^32^. The analyses were adjusted for age, sex, and the first 10 genetic principal components, and, in FinnGen, also for the genotyping batch. In the sex-stratified analyses, sex was not included as a covariate.

The METAL software (v.2011-03-25)^33^ was used to do inverse-variance weighted fixed-effect meta-analyses of the GWAS results from FinnGen and the Estonian Biobank. Variant data from the Estonian biobank were converted from hg19 to hg38 prior to the meta-analysis.

### Characterization of the association signals

We defined a locus as a window of 1 MB (± 500,000 bases from the association lead variant) containing at least one variant associated with SNHL at P<5×10^−8^. By manually annotating every gene within the loci and prioritizing those according to a relevant biological function using literature and database resources such as Genbank^34^ and Uniprot^35^, we were able to identify potential candidate genes for the loci that had not been linked to SNHL in previous studies.

### Functional annotations

We employed MAGMA v1.10 (linux)^36^ for conducting gene-based and gene-set analysis on SNHL data. The input file utilized in the analysis comprised meta-analyzed results from the SNHL GWAS conducted in the general population. The 1000 Genomes project European population was used as a reference sample for LD corrections. The variants were mapped to gene locations according to human genome build 38 (NCBI38.gene.loc). To better capture variants in the regulatory regions, we added a 35kb window upstream and a 10kb window downstream of the genes. Subsequently, the association signals of the variants were aggregated to gene-level statistics.. The gene-set level analysis used the 2023.2 version of the MSigDB gene set file, comprising 34,494 curated gene sets and gene ontology (GO) terms.

### eQTL colocalizations

Gene expression and observed SNHL associations were colocalized using the “coloc” R-library function ‘coloc.abf’^37^. For every SNHL-associated locus, colocalization was evaluated for every gene within a 1MB window from the association lead variant. From the GTExportal (https://www.gtexportal.org/home/datasets), we downloaded variant-gene expression associations (GTEx v8, accessed on 19/05/2022) for the analysis. SNHL-relevant tissues were used in the analysis, including whole blood, brain (cerebellum, cortex, and hippocampus), adipose (subcutaneous, visceral), and tibial nerve. We utilised ≥ 0.8 posterior probabilities as a limit for significant results^38^.

### Genetic correlations

Genetic correlations were computed using the LDSC software^39^. Genetic correlations between SNHL and 767 other traits were computed using data taken from the MRC Integrative Epidemiology Unit (IEU)’s GWAS database (https://gwas.mrcieu.ac.uk/). The significance limit for correlations was set at a P_FDR_ < 0.05.

### Subgroup effect differences

We estimated if there were significant differences in the effect sizes of the lead variants at each locus between the analytical subgroups (i.e., those diagnosed before the age of 55 years vs. those diagnosed at or after 55 years of age, and females vs. males) using the following formula, with the corresponding P-values calculated from the normal distribution:

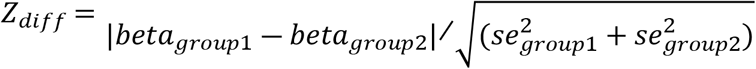

Effect size differences with P_FDR_<0.05 were deemed statistically significant.

We used five prediction tools for assessing the consequences of the missense mutations (Table 2). PolyPhen-2^40,41^, Sorting Tolerant From Intolerant (SIFT)^42,43^, and Protein Variation Effect Analyzer (PROVEAN)^44^ assess the consequences of the mutation using conservation-based prediction, evaluating the variants by comparing similar proteins and considering local sequence conservation. To utilize meta-prediction and machine learning, we used the Rare Exome Variant Ensemble Learner (REVEL)^45^, which has been shown to improve the prediction accuracy compared to methods based on sequence conservation alone. OpenCRAVAT interface was utilized in this approach (https://www.opencravat.org/), and we also report the consensus among all the missense prediction tools included in that platform. We exploited AlphaMissense, which combines structural context and evolutionary conservation of the mutated position to predict the consequences of missense variants^46,47^.

## Results

22 loci associated with SNHL in general population

In our genome-wide meta-analyses of SNHL, we discovered 22 genome-wide significant (P<5×10^−8^) loci associated with SNHL (Figure 1A, Table S2). Of these, seven loci had not been reported in association with SNHL in prior studies (Table 1, regional plots Figures S1-S7), whereas the remaining 15 were located proximally to known SNHL risk loci (Table S2)^13,18,19,21,23,25,26^. The association signals at 6p21.31-6p22.2 located in the HLA region extend beyond the 1 Mb locus definition and were counted as one locus. Although the FinnGen and the Estonian Biobank results were generally consistent, we observed some heterogeneity in the complex associations at the HLA and 19q13.33 loci (Table S2-S6).

**Table 1.**
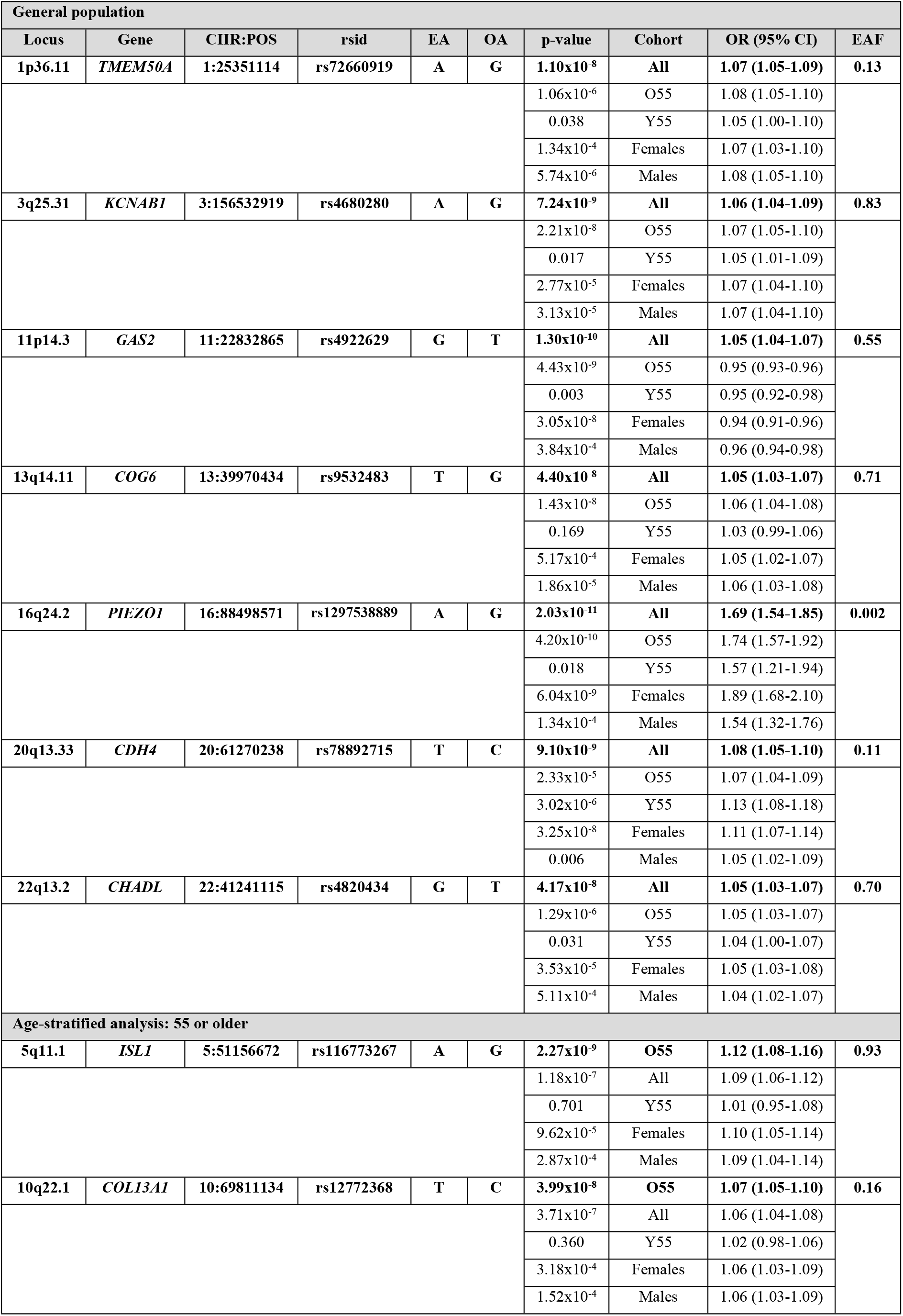
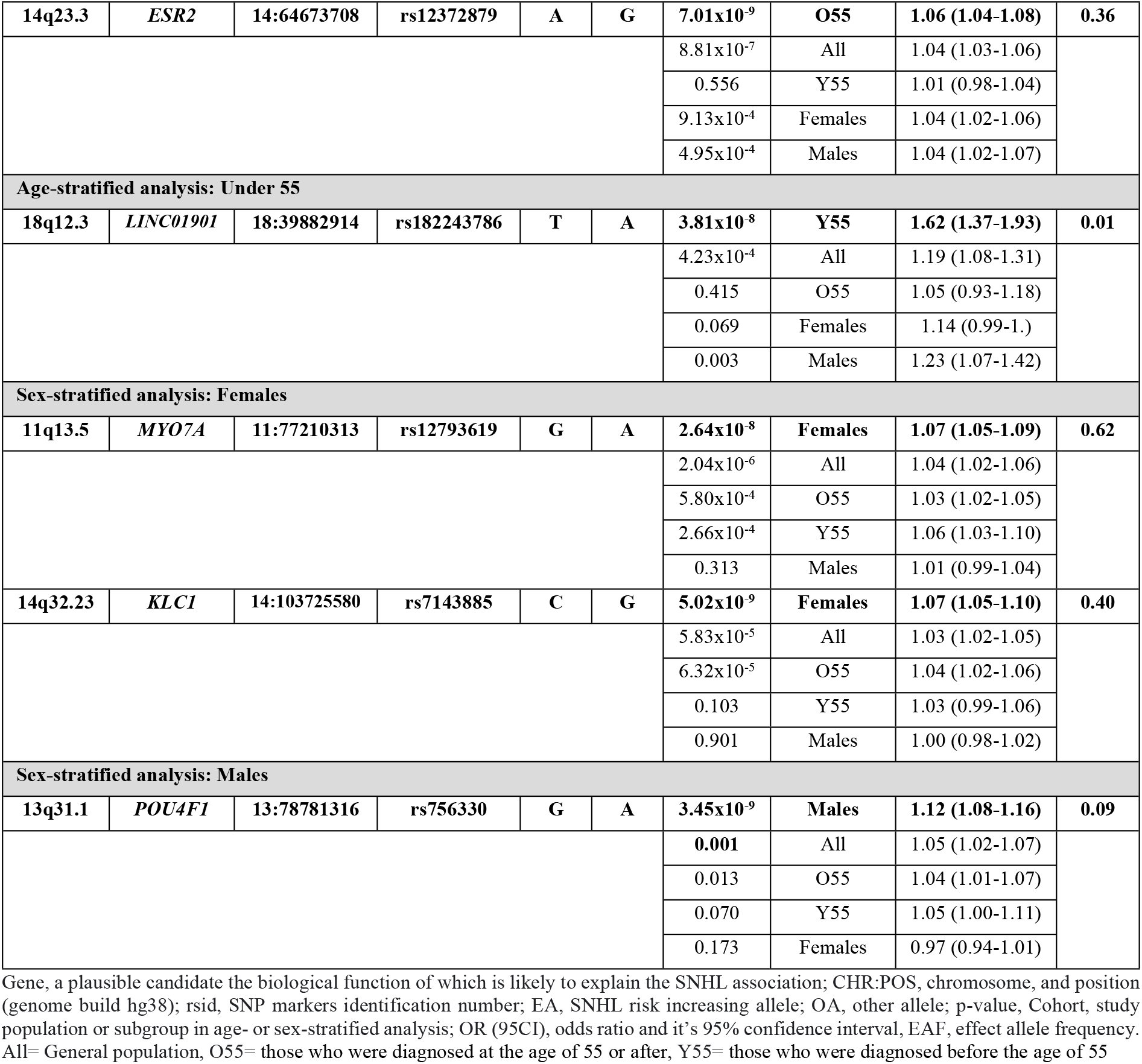
Novel SNHL-associated genome-wide significant (P<5×10^−8^) loci identified in the genome-wide meta-analyses based on FinnGen and the Estonian Biobank. A full list of SNHL-associated genome-wide significant loci identified in the genome-wide meta-analyses can be seen in Table S2-Table S6.

**Figure 1.**
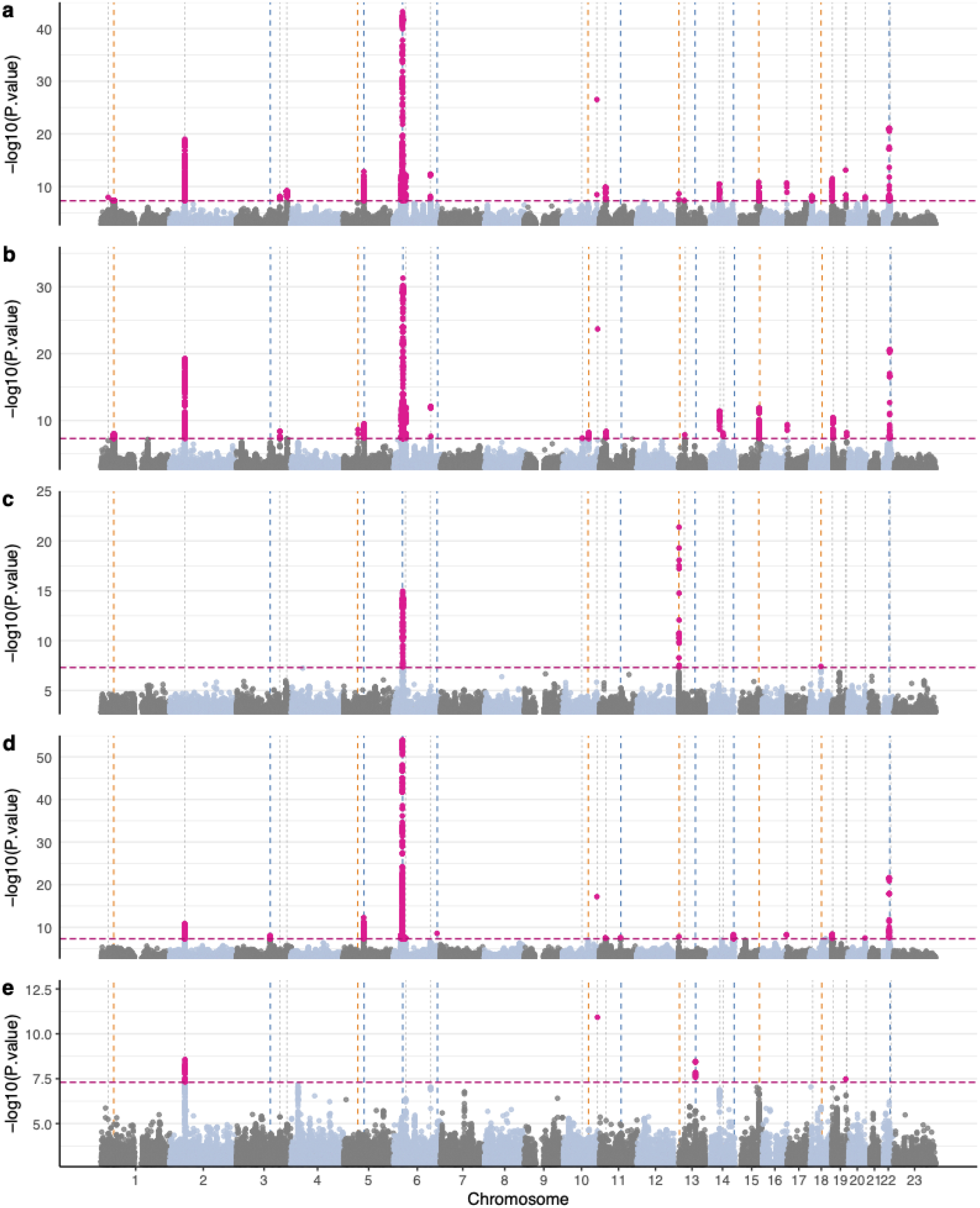
Manhattan plots of the SNHL associations in a) the general population, b) 55 years or older, c) younger than 55, d) females, and e) males. Vertical dashed lines indicate loci that were associated with SNHL with genome-wide significance (P<5×10^−8^) in at least one of the analytical subgroups; color coding is as follows: orange=FDR-significant effect size difference in o55 vs. y55; blue=pFDR-significant effect size difference in females vs. males; gray=no FDR-significant effect size differences.

### Age-specific results

We identified 20 genome-wide significant (P<5×10^−8^) loci within the subgroup of SNHL cases diagnosed at the age of 55 or older, with three of these loci not previously linked to SNHL (Figure 1B, Table 1, Figure S8-S10, Table S3). In the subgroup of SNHL cases diagnosed before the age of 55, we observed three genome-wide significant loci of which one was novel (Figure 1C, Table 1, Figure S11, Table S4). Two of the 15 genome-wide significant loci observed in the female subgroup have not previously been associated with SNHL (Figure 1D, Table 1, Figure S12-S13, Table S5). Four genome-wide significant loci were identified from the male subgroup; one was novel (Figure 1E, Table 1, Figure S14, Table S6).

### Effect differences

We observed significant age differences in effect sizes for lead variants near *CCDC17* (*coiled-coil domain containing 17*), *ISL1* (*ISL LIM homeobox 1*), *EXOC6* (*exocyst complex component 6*), *GJB2* (*gap junction protein beta 2*), *ISG20* (*interferon stimulated exonuclease gene 20*) and 18q12.3 locus, rs182243786, which was classified as *LINC01901*(*long intergenic non-protein coding RNA 1901*)(Figure 2A). Additionally, significant sex differences in effect sizes were observed for lead variants near *ILDR1* (*immunoglobulin like domain containing receptor 1*), *ARHGEF28* (*Rho guanine nucleotide exchange factor 28*), *HLA* rs192376296, *SYNJ2* (*synaptojanin 2*), *MYO7A* (*myosin VIIA*), *POU4F1* (*POU class 4 homeobox 1*), *KLC1* (*kinesin light chain 1*) and *TRIOBP* (*TRIO and F-actin binding protein*) (Figure 2B).

**Figure 2.**
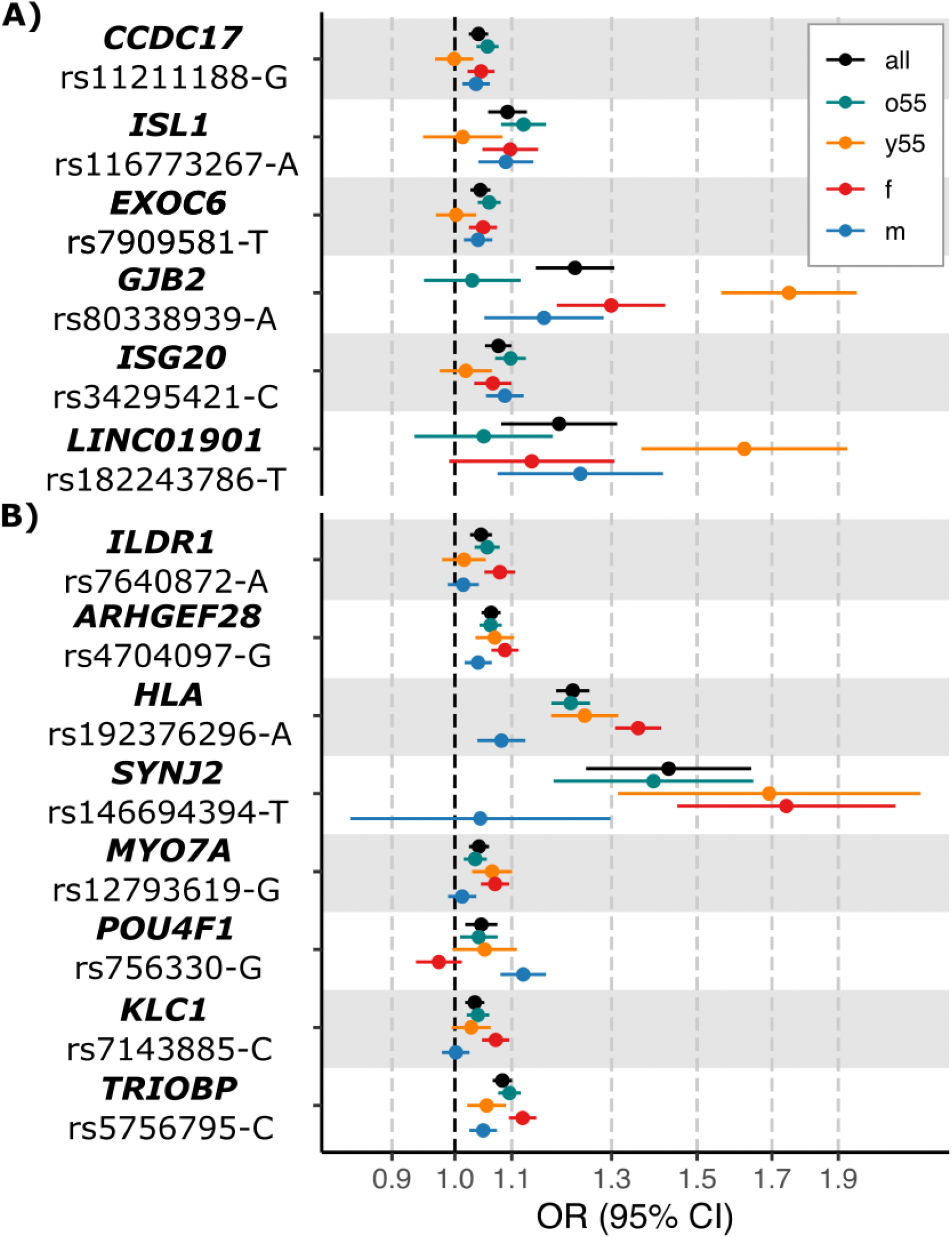
Significant differences in the effect sizes of the lead variants at the SNHL-associated loci between (A) the age-of-onset-stratified subgroups and (B) sex-stratified subgroups. Variants are identified by candidate gene, rsid, and SNHL risk-increasing allele. General population [all, black]; older than 55 years [o55, green]; younger than 55 years [y55, orange]; females [f, red]; males [m, blue].

### MAGMA & eQTL colocalizations

Overall, we identified 80 genes significantly associated with SNHL in the MAGMA gene-based test (P<2.52e-6 to correct for 19,823 genes; Table S7). The MAGMA gene-set analysis revealed the most significant enrichments in the pathway related to the stereocilium base (P=1.01×10^−8^, Figure 3). Analysis also revealed pathways regulating the differentiation of inner ear auditory receptor cells (P=1.42×10^−7^, Figure 3) and hair cells (P=8.72×10^−7^, Figure 3). A fourth, significant enrichment was observed for the mechanoreceptor differentiation pathway (P=1.00×10^−6^, Figure 3). Colocalizations between SNHL GWAS and gene expression signals (eQTL) were observed in every tissue studied (Figure S16). The expression of the 102 genes and SNHL signals was found to colocalize in at least one tissue (posterior probability [PP] for the shared variant ≥ 0.8). For example, SNHL association signals and expression of *CCDC163* (*CCDC163 homolog*) and *SPTBN1* (*spectrin beta, non-erythrocytic 1*) were colocalized in six out of the seven tissues under investigation (Figure S16).

**Figure 3.**
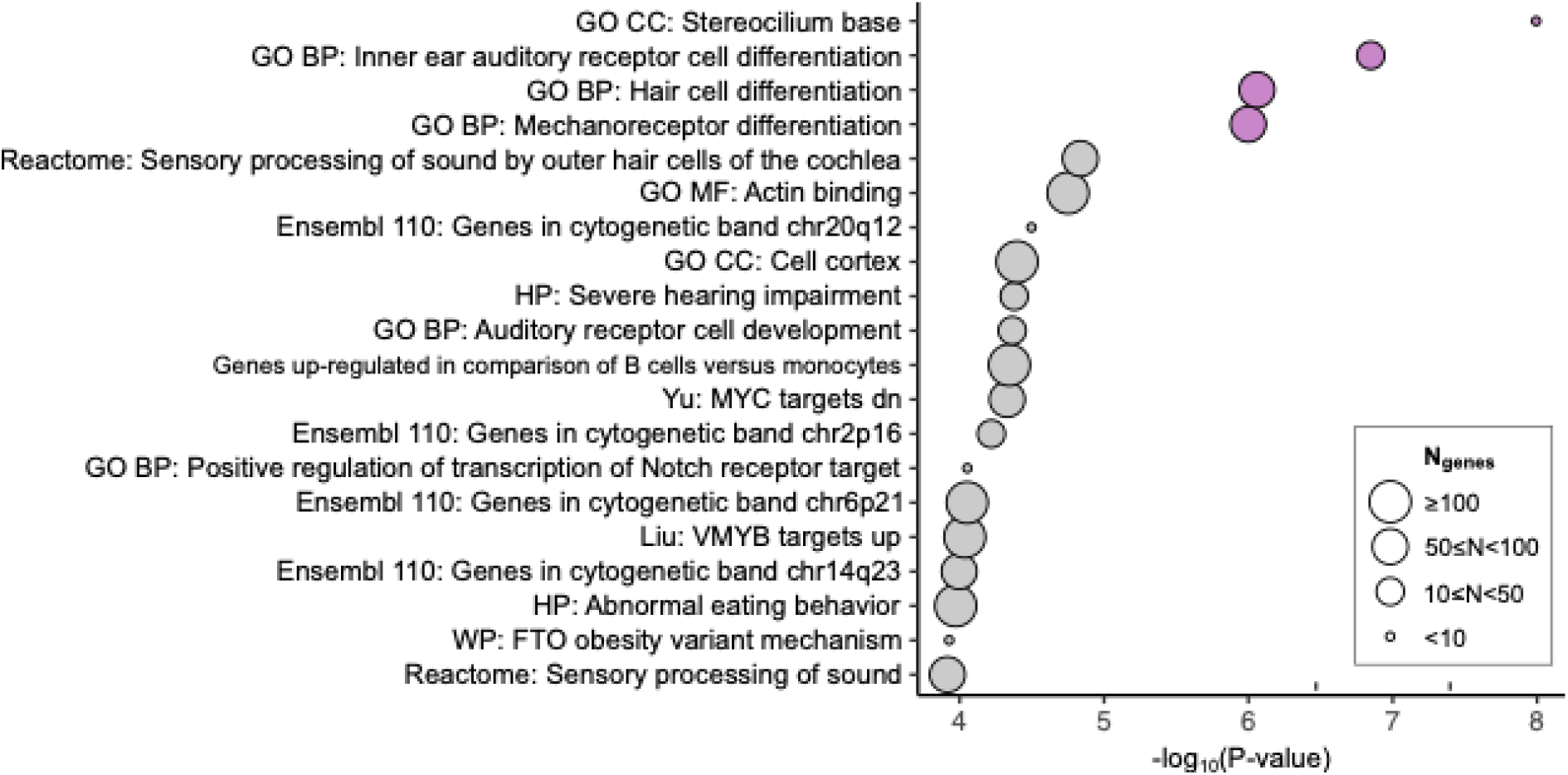
MAGMA gene-set analysis. We conducted MAGMA gene-set analysis using 34,494 curated gene sets and Gene Ontology (GO) terms from the MSigDB^48^ (version 2023.3). The plot displays the 20 most significant gene sets, with four highlighted in pink, indicating significance following false discovery rate (FDR) correction.

### Genetic correlation analysis reveals extensive links between SNHL and hearing-related traits

We tested for genetic correlations of SNHL with 767 other traits. The strongest positive correlations between SNHL and other traits were observed with ‘hearing difficulty/problems: Yes’ (rg=0.68, Figure 4) and ‘hearing aid user’ (rg=0.63, Figure 4). Also, we observed positive correlations with other hearing-related traits such as ‘hearing difficulty/problems with background noise’ (rg=0.52, Figure 4) and ‘tinnitus: Yes, now most or all of the time’ (rg=0.42, Figure 4). Interestingly, a negative correlation was observed with ‘tinnitus: No, never’ endpoint (rg=-0.22, Figure 4). Additionally, a strong negative correlation that reached the significance threshold was observed with ‘H40: Glaucoma’ (ukb-d-H40, rg=-0.54), but since the result did not replicate with another glaucoma endpoint (bbj-a-121, rg=-0.02), this result is possibly a false positive (Table S8).

**Figure 4.**
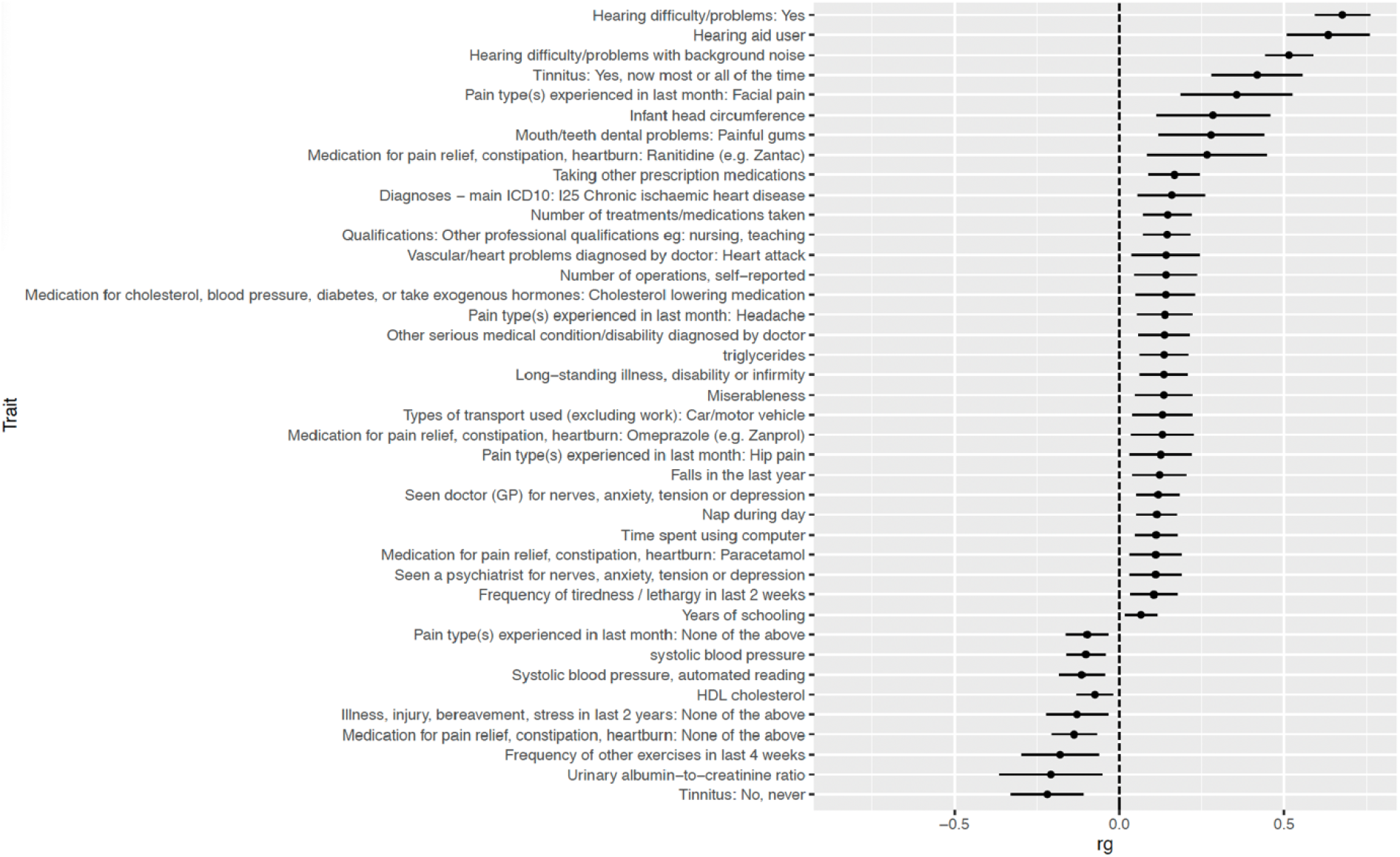
Genetic correlations between SNHL and other traits. Genetic correlations were calculated using LDSC-software^39^, SNHL summary statistics from the general population, and for other traits, data were extracted from the GWAS database provided by the MRC Integrative Epidemiology Unit (IEU) (https://gwas.mrcieu.ac.uk/). rg, genetic correlation coefficient value. Figure 4 only displays the significant correlations (false discovery rate-corrected p-value [pFDR]<0.05); Table S8 displays the genetic correlations between SNHL and all examined traits.

In the prediction of the consequences for missense variants, *SYNJ2* variant rs146694394 indicated the most evident influence on protein structure-function (Table 2). For *CRIP3* rs2242416, the predictions were somewhat dependent on the exact deviation in the sequence, all of the variant types showing rather mild effects. Similarly, while modest, the *TRIOBP* variant rs5756795 showed mild effects on protein. AlphaMissense predictions correlated poorly with the other prediction results.

**Table 2.** Prediction of the consequences of the missense variants with selected algorithms. The prediction and scoring are provided for all the predictors. Unfortunately, AlphaMissense prediction was not available for rs2242416. Consensus score obtained from OpenCRAVAT is reported, where the number refers to predictions reporting the mutation being benign or tolerated; the total number of predictors applied is shown as well.

| rsid/Gene | Mutation | SIFT |  | PolyPhen-2 HDIV |  | PROVEAN |  | REVEL |  | CRAVAT |  | AlphaMissense |  |
| --- | --- | --- | --- | --- | --- | --- | --- | --- | --- | --- | --- | --- | --- |
| rs146694394<br>*SYNJ2* | T656M | Damaging | 0.001 | Probably damaging | 1.000 | Deleterious | -5.504 | Uncertain | 0.655 | 2/13 |  | Likely pathogenic | 0.730 |
| rs2242416<br>*CRIP3* | I188S | Tolerated | 0.397 | Possibly damaging | 0.793 | Neutral | -0.060 | Uncertain | 0.473 | 4/11 |  | N/A | N/A |
|  | I188T | Tolerated | 0.397 | Benign | 0.093 | Neutral | 1.39 | Uncertain | 0.314 | 8/11 |  | N/A | N/A |
|  | I188N | Tolerated | 0.304 | Probably damaging | 0.982 | Neutral | -0.870 | Uncertain | 0.598 | 3/11 |  | N/A | N/A |
| rs5756795<br>*TRIOBP* | F1187I | Damaging | 0.023 | Possibly damaging | 0.678 | Neutral | -2.15 | Benign | 0.102 | 8/11 |  | Likely benign | 0.258 |
|  | F1187L | Tolerated | 0.089 | Benign | 0.006 | Neutral | -2.02 | Benign | 0.042 | 11/11 |  | Likely pathogenic | 0.745 |

## Discussion

Twenty-two SNHL-associated loci were associated in the genome-wide meta-analysis of the general population (35,960 cases and 531,059 controls), seven of which were previously unreported. Furthermore, three previously unreported loci were observed in the sex-stratified analyses and four in the age-stratified analyses, totaling 14 previously unreported SNHL risk loci observed in this study. We also replicated multiple loci that have previously been associated with SNHL, for example, those harboring *SYNJ2* (*synaptojanin 2*), *TRIOBP* (*TRIO and F-actin binding protein*), and *GJB2* (*gap junction protein beta 2*) that are well-known SNHL risk loci^25^.

### Previously unreported SNHL risk loci

We detected previously unreported loci near genes that play a role in mechanotransduction, such as *MYO7A* (*myosin VIIA*) and *PIEZO1* (*piezo type mechanosensitive ion channel component 1*). In previous functional studies, *MYO7A* has been observed to play a role in mechanotransduction, and it is expressed in mouse cochlea^49^. It has also been linked to hearing loss in previous candidate gene studies^50,51^, but our genome-wide meta-analysis was the first to report its association with SNHL. Also, *PIEZO1* is expressed in inner and outer hair cells and is proposed to have a role in mechanotransduction^52^. Since mechanotransduction transforms sound energy into neuronal impulses, it is a crucial process in hearing^53^, and potential defects in this process may contribute to the pathophysiology of SNHL. In addition, we observed previously unreported SNHL associations near *ISL1* (*ISL LIM homeobox 1*) and *POU4F1* (*POU class 4 homeobox 1*) that potentially regulate differentiation and maintenance of inner ear sensory neurons^54^. Also, locus near *GAS2* (*growth arrest specific 2*) was associated with SNHL. According to earlier research, *GAS2* preserves the integrity of the microtubule bundle in the cochlear supporting cells, and any defects in its expression could compromise the cochlea’s ability to transmit sound energy^55^.

### Functional annotations & effect differences

Several of the same genes that we had annotated to be potential candidate genes were identified in the MAGMA gene-based analysis, supporting our interpretation. In the MAGMA gene-set analysis, enrichments were observed in the pathways related to the structure of the stereocilia and its differentiation, such as stereocilium base and inner ear auditory receptor cell differentiation. These results were in line with previously reported results, which have reported similar enrichments for different gene sets related to the structures of the inner ear^23,25^. We observed genetic correlations between a number of different hearing-related traits, which indicates that these traits share a significant amount of genetic architecture. Also, these results were strongly in line with previously reported genetic correlations^18^. Based on consequence prediction performed for missense variants, it is likely that none of the variants cause complete inactivation of the target protein. Effect sizes were significantly larger in SNHL cases diagnosed before the age of 55 for lead variants near *GJB2* and *LINC01901*, while effects of lead variants in/near *CCDC17, ISL1, ISG20*, and *EXOC6* were larger in those diagnosed at the age of 55 or older. The sex-stratified comparisons indicate that the effect sizes for SNHL lead variants in/near *ILDR1, ARHGEF28, HLA, SYNJ2, MYO7A, KLC1*, and *TRIOBP* were significantly larger in females compared with males. Only at the *POU4F1* locus was the lead variant effect size significantly larger in males than in females.

## Limitations & Conclusions

We have only employed computational methods and, thus, our findings would benefit from further functional analyses to identify truly causal genes. The eQTL colocalization analysis lacks sensory tissue-related data, such as cochlea-specific datasets, which would be relevant to this study. Replicating these findings in diverse ethnic groups would enhance generalizability, as our sample is limited to European ancestry, particularly the distinct Finnish gene pool.

In summary, we were able to advance our knowledge of the genetic basis of SNHL by discovering a considerable amount of previously unreported loci and carrying out numerous downstream analyses. Interestingly, we observed associations near genes that could affect SNHL pathogenesis through the mechanotransduction process or compromise the functions other inner ear structures. The involvement of biological pathways related to the structure of inner ear in the pathogenesis of SNHL was validated in downstream analyses. Age- and sex-stratified approach provided additional insights into the genetic factors of SNHL. Our work helps us to better understand the genetic causes of SNHL and could pave the way for future studies into novel therapies and preventative strategies.

## Data Availability

Available after publication GWAs catalog (not yet) and upon request

## Statements and Declarations

We want to acknowledge the participants and investigators of FinnGen study. The FinnGen project is funded by two grants from Business Finland (HUS 4685/31/2016 and UH 4386/31/2016) and the following industry partners: AbbVie Inc., AstraZeneca UK Ltd, Biogen MA Inc., Bristol Myers Squibb (and Celgene Corporation & Celgene International II Sàrl), Genentech Inc., Merck Sharp & Dohme LCC, Pfizer Inc., GlaxoSmithKline Intellectual Property Development Ltd., Sanofi US Services Inc., Maze Therapeutics Inc., Janssen Biotech Inc, Novartis AG, and Boehringer Ingelheim International GmbH. Following biobanks are acknowledged for delivering biobank samples to FinnGen: Auria Biobank (www.auria.fi/biopankki), THL Biobank (www.thl.fi/biobank), Helsinki Biobank (www.helsinginbiopankki.fi), Biobank Borealis of Northern Finland (https://www.ppshp.fi/Tutkimus-ja-opetus/Biopankki/Pages/Biobank-Borealis-briefly-in-English.aspx), Finnish Clinical Biobank Tampere (www.tays.fi/en-US/Research_and_development/Finnish_Clinical_Biobank_Tampere), Biobank of Eastern Finland (www.ita-suomenbiopankki.fi/en), Central Finland Biobank (www.ksshp.fi/fi-FI/Potilaalle/Biopankki), Finnish Red Cross Blood Service Biobank (www.veripalvelu.fi/verenluovutus/biopankkitoiminta), Terveystalo Biobank (www.terveystalo.com/fi/Yritystietoa/Terveystalo-Biopankki/Biopankki/) and Arctic Biobank (https://www.oulu.fi/en/university/faculties-and-units/faculty-medicine/northern-finland-birth-cohorts-and-arctic-biobank). All Finnish Biobanks are members of BBMRI.fi infrastructure (www.bbmri.fi). Finnish Biobank Cooperative -FINBB (https://finbb.fi/) is the coordinator of BBMRI-ERIC operations in Finland. The Finnish biobank data can be accessed through the Fingenious® services (https://site.fingenious.fi/en/) managed by FINBB. We want to acknowledge the participants and staff of the Estonian Biobank for their contributions. The Estonian Genome Centre analyses were carried out in part in the High Performance Computing Center, University of Tartu. This study was conducted using the Estonian Center of Genomics/Roadmap II funded by the Estonian Research Council (project number TT17).

